# Endometriosis and Early-Onset Cardiovascular Disease Across Levels of Systemic Inflammation: A Cross-Sectional Study of NHANES 1999–2006

**DOI:** 10.64898/2026.09.29.26364168

**Authors:** Chidinma Oli, Natalie Chichetto

## Abstract

**Background:** Endometriosis is a chronic gynecologic condition which has been linked with higher cardiovascular disease (CVD) risk. Although systemic inflammation is central to endometriosis and CVD pathophysiology, it remains unclear whether endometriosis is associated specifically with early-onset CVD and whether systemic inflammation modifies this association.

**Methods:** This was a cross-sectional study of 5,345 women aged 20–54 years from the National Health and Nutrition Examination Survey (1999–2006). Endometriosis and early-onset CVD were self-reported. Early-onset CVD was defined as a composite measure of coronary heart disease, angina/angina pectoris, heart attack, congestive heart failure, or stroke. Systemic inflammation was measured using C-reactive protein [CRP; low/moderate (≤3 mg/L) and high (>3mg/L)] and the systemic immune-inflammation index (SII; tertiles). Survey-weighted logistic regression was used to assess associations and heterogeneity by systemic inflammation, with stabilized inverse probability weighting (IPW) to adjust for confounding.

**Results:** Of the 5,435 women, 370 (6.9%) reported an endometriosis diagnosis and 162 (3.0%) early-onset CVD. After IPW adjustment for age, body mass index, smoking status, education, age at menarche, physical activity and race/ethnicity, endometriosis was associated with higher odds of early-onset CVD (prevalence odds ratio [POR] = 1.91, 95% CI: 1.23–2.97). This association was strongest among women with high CRP (POR= 2.33, 95% CI: 1.49–3.66), but not among those with low/moderate CRP levels (p-interaction= 0.086). Across SII strata, the strongest association was observed among women in the medium SII tertile (POR = 2.92, 95% CI: 1.47–5.78), with no associations in the low or high tertiles (p-interaction = 0.288).

**Conclusion:** Endometriosis was associated with higher odds of early-onset CVD in this nationally representative sample of U.S. women. Although the strength of the association varied across CRP and SII strata, formal tests for interaction were not statistically significant. These findings should be understood as exploratory and require further investigation in prospective studies

## Introduction

Cardiovascular disease (CVD) is the leading cause of mortality in women globally and in the United States [1, 2], yet younger and middle-aged women are often underrecognized in risk assessment and prevention efforts. This gap has contributed to delayed diagnoses, missed opportunities for intervention, and a rising burden of early-onset CVD (i.e., cardiovascular events occurring before age 65) in women [3]. Early-onset CVD has substantial implications including higher rates of clinical comorbidities (e.g., metabolic syndrome, diabetes, and obesity), increased cumulative healthcare costs and an elevated lifetime risk of recurrent cardiovascular events [3–7].

Notably, women account for nearly half of incident cases of early-onset CVD despite generally lower prevalence of conventional cardiovascular risk factors [8]. This pattern may reflect two factors. First, these conventional risk factors (e.g., diabetes, smoking, and hypertension) appear to have stronger harmful effects such as an increased risk of CVD in women, compared to men with similar risk factors [9]. Specifically, diabetes was associated with a 3-7 times higher risk of CVD among women compared with a 2-3 times higher risk in men [10, 11]. Further, smoking increases the relative risk of CVD in women by 25–55% despite lower smoking prevalence [12]. Second, growing evidence underscores the role of sex-specific, gynecologic conditions such as endometriosis in shaping CVD risk [13–15].

Endometriosis is a chronic, estrogen-dependent inflammatory condition affecting approximately 5-10% of reproductive-aged women [13, 14, 16, 17]. It is defined by the growth of endometrial-like ectopic tissues and stroma outside the uterine cavity and primarily inside the ovaries and pelvic peritoneum [17, 18]. Signs and symptoms include chronic pelvic pain, dysmenorrhea, dyspareunia, dyschezia (pain during defecation) and reduced fertility [19, 20].

Shared inflammatory and immune pathways are hypothesized as a leading mechanism that may explain the association between endometriosis and CVD [21–24]. The endometrial ectopic tissues recruit and activate immune cells like macrophages, lymphocytes and neutrophils, which trigger the release of inflammatory markers such as C-reactive protein (CRP). [25–27]. At elevated levels, these inflammatory markers may exacerbate chronic inflammatory processes and affect the mechanism of action of endothelial nitric oxide synthase, which results in endothelial cell dysfunction (i.e., damage to the vascular endothelium), a key process implicated in atherosclerosis and the development of CVD [26, 28–30].

Since endometriosis is typically diagnosed during the reproductive years [31], women with endometriosis may experience prolonged exposure to chronic systemic inflammation for decades before cardiovascular risk is typically assessed, potentially contributing to an earlier-onset of CVD. However, most studies have examined overall CVD risk without considering whether endometriosis contributes to an earlier onset of CVD [32, 33]. Additionally, although chronic inflammation has been documented in endometriosis [22, 34], the extent to which it may modify the association between endometriosis and early-onset CVD has not been formally tested.

Therefore, our study aims to examine the association between endometriosis and early-onset CVD and assess whether it differs across levels of systemic inflammation measured by CRP and the systemic immune-inflammation index (SII), using data from the National Health and Nutrition Examination Survey (NHANES). Clarifying this heterogeneity may help identify women with a higher burden of early-onset CVD. We examined both markers because while elevated levels of CRP may signify chronic low-grade inflammation characteristic of chronic condition like endometriosis, [35, 36], it may not fully capture the complex immune-inflammatory interactions that occur in endometriosis [37]. In contrast, SII is a composite measure that integrates the inflammatory, immune activation, and thrombotic mechanisms that are central to the pathogenesis of both endometriosis and CVD [37, 38], making it a complementary and biologically meaningful measure.

## 2.0 Methods

### Study Design and Settings

This is a cross-sectional study using data from the 1999-2006 cycles of the National Health and Nutrition Examination Survey (NHANES) [39]. These survey cycles were selected because they included publicly available data on self-reported endometriosis diagnosis (https://wwwn.cdc.gov/nchs/nhanes/). NHANES is administered by the National Center for Health Statistics (NCHS) and collects person-level demographic, health, nutrition and disease status information of the civilian, non-institutionalized U.S. population at two-year intervals, using a complex stratified, clustered, and four-stage sample design[39]. Data are collected through household interviews, physical examination and laboratory measures at a mobile examination center (MEC) [39]. The study was approved by the NCHS ethics review board, and all participants provided written informed consents prior to enrollment.

### Study Population

Our study comprised of 41,474 participants from the 1999–2006 NHANES survey cycles, of which 21,210 were female. NHANES administers the reproductive health questionnaire to females aged ≥12 years (n = 15,439) at the mobile examination center (MEC) using the computer-assisted personal interviewing system. Of these, 6,508 were aged 20-54 years and 5,575 provided a response to self-report endometriosis (including “don’t know/refused”). After excluding participants who responded “don’t know” on self-report endometriosis diagnosis (n=18) and those with missing CVD or covariate data, the final analytic sample included 5,345 women (Figure 1).

**Figure 1:**
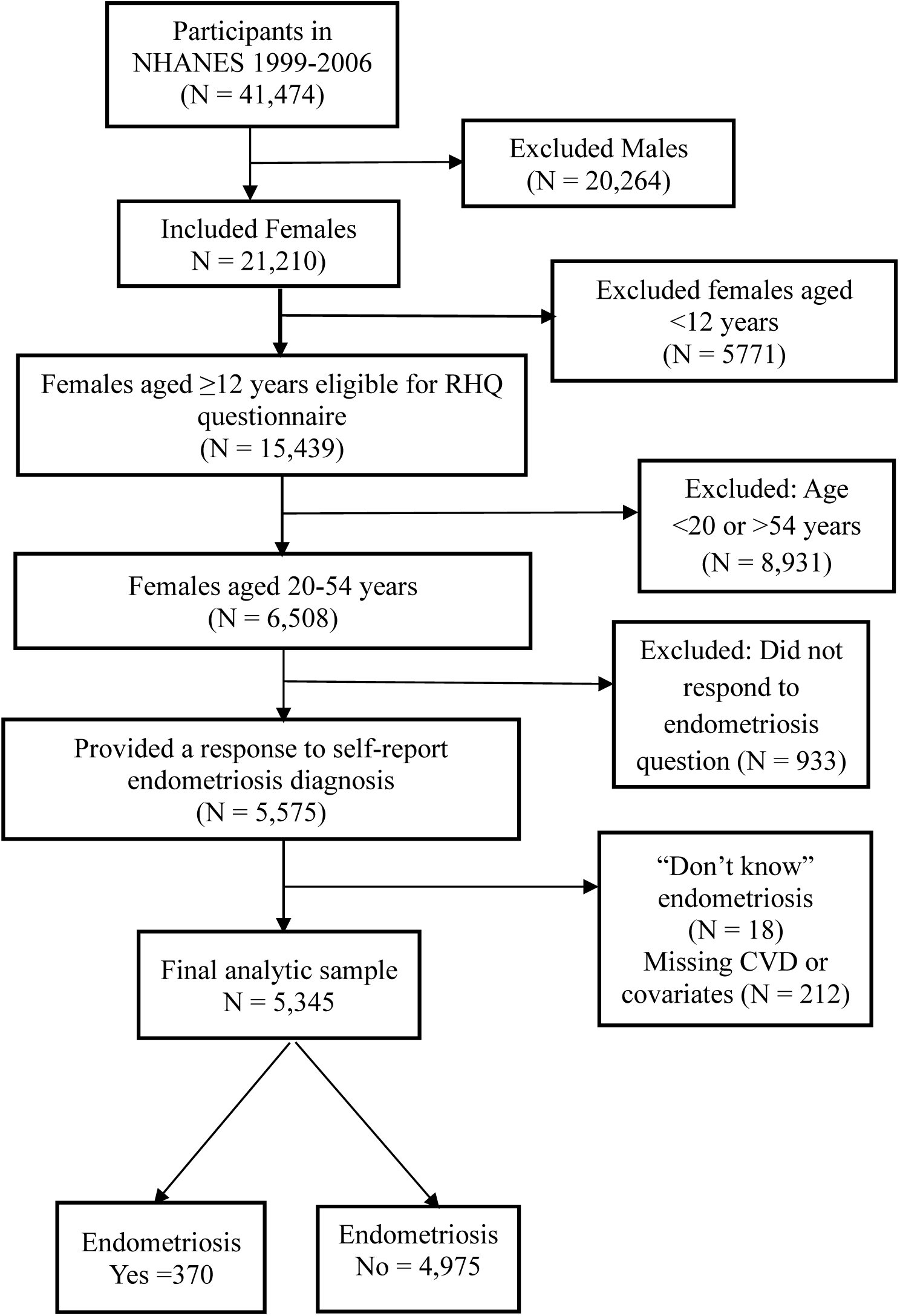
Flow chart of participant selection in NHANES 1999–2006 for U.S women aged 1996-2006.

## Measures

### Exposure Measure

The exposure measure was self-report of endometriosis diagnosis by a physician or health professional via the following survey item on the reproductive health questionnaire: *“Has a doctor or other health professional ever told that you had endometriosis?*” Any woman who self-reported “Yes” to the question was considered to have endometriosis and those who responded “No” were not. With an overall confirmation proportion of 84% (range: 72% to 95%) there is evidence to suggest that self-report endometriosis diagnosis is reasonably valid [40].

### Outcome Measure

The outcome measure was early-onset CVD defined as a self-reported physician diagnosis of any of the following conditions: coronary heart disease, angina/angina pectoris, heart attack, congestive heart failure, or stroke before age 55. Early-onset CVD status was determined using response from the survey items on the medical conditions’ questionnaire, which asked participants, *“Has a doctor or other health professional ever told you that you had (specific condition - e.g., coronary heart disease*)? Participants who responded “yes” to at least one condition were classified as having CVD, and those who responded “no” to all were classified as not having CVD. Self-report CVD has demonstrated good validity in prior work [41, 42].

### Effect Modifier

The potential effect modifier of interest was systemic inflammation, assessed using two complementary biomarkers: CRP and SII. Both biomarkers were assessed from blood samples collected during the mobile examination visit using a standardized protocol [43]. CRP was quantified using high-sensitivity latex-enhanced nephelometry. Due to sparse cell counts of early-onset CVD events, CRP was categorized as low/moderate (≤3 mg/L) and high (>3 mg/L) to improve model stability while maintaining clinically meaningful cut points [44, 45]. SII was calculated as platelet count × neutrophil count / lymphocyte count [38]. Hematology testing is performed on blood specimens collected from participants and evaluated for neutrophil (10³ cells/µL), segmented neutrophil (10³ cells/µL), and platelet (10³ cells/µL) counts. Cell counts are determined using the Coulter MAXMs method (Beckman Coulter, Miami, FL, USA) [43]. Values outside physiologically plausible ranges for lymphocyte (n=1) and platelet counts (n=2) were treated as missing (lymphocytes >40 ×10³ cells/µL; platelets <50 or ≥999 ×10³ cells/µL), affecting fewer than 1% of observations. SII was divided into three survey-weighted tertiles: low (<33rd percentile), medium (33rd–67th), and high (>67th percentile).

### Assessment of covariates

Covariate selection was informed by a directed acyclic graph (DAG) (Figure 2) constructed based on prior literature to identify a minimal sufficient adjustment set. Selected covariates represent factors associated with both endometriosis and CVD risk and included age, race/ethnicity, body mass index (BMI), smoking status, education, insurance status, age at menarche, and physical activity [46–50].

**Figure 2:**
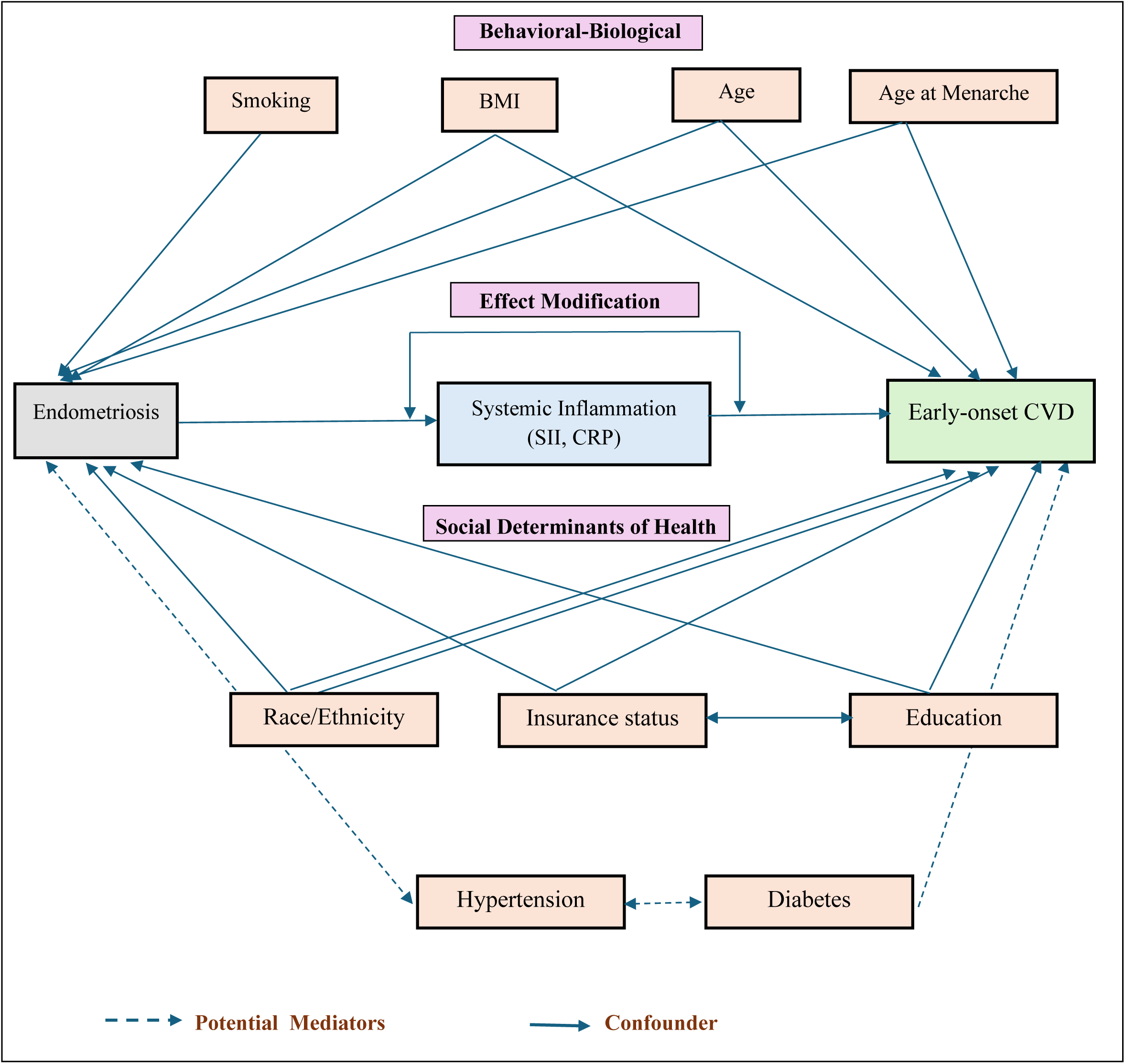
Directed Acyclic Graph.

Age (years), age at menarche (years) and BMI (kg/m^2^), calculated from measured height and weight were treated as continuous variables. Smoking status was categorized into current, former and never smokers. Participants’ educational level was categorized as less than high school, high school, or college and above. For insurance, participants were classified as “Insured” if they reported having any form of health insurance and “Uninsured” if they reported no insurance. Race/ethnicity (non-Hispanic white, non-Hispanic black, Hispanic, and Others). Physical activity was measured using a survey item asking participants to select the description that best reflected their usual daily activity level. Responses were grouped into three categories: low (mostly sitting), moderate (standing or walking frequently), and high (lifting loads, climbing stairs/hills, or performing heavy work). This item has been validated against accelerometry counts and used in prior NHANES studies [51, 52].

### Statistical Analysis

To account for the complex, multistage sampling design of the NHANES, we conducted all the statistical analyses with the appropriate sampling weights [39]. Since both the reproductive health questionnaire items and laboratory data were collected during the mobile examination visit, we used the recommended MEC weights consistent with National Center for Health Statistics (NCHS) analytic guidelines [53]. To account for pooling multiple survey cycles (1999–2006), 4-year MEC weights (WTMEC4YR) for the 1999–2002 cycles were divided by two, and 2-year MEC (WTMEC2YR) weights for 2003–2006 were divided by four to create an 8-year combined MEC weight, following NCHS analytic guidelines [53].

Descriptive statistics were used to compare characteristics between women with and without self-report endometriosis. We used survey-weighted means and standard errors to summarize continuous variables for approximately normally distributed variables, while non-normal distributions were summarized as medians and interquartile ranges (IQRs). Categorical variables were summarized using weighted frequencies and percentages. Group differences by self-report endometriosis diagnosis was assessed using weighted chi-square test for categorical variables and weighted t-tests or Wilcoxon rank-sum tests for normally distributed variables and for skewed continuous variables respectively.

To address our study aims and reduce bias due to measured confounding, stabilized inverse probability of weighting (IPW) was used to balance measured covariates between women with and without endometriosis [54, 55]. IPW was chosen because early-onset CVD is a relatively rare outcome and the number of covariates was large relative to the number of events, conditions under which propensity-based methods such as IPW have been shown to yield more stable, less biased, and more robust estimates than conventional regression adjustment [56, 57]. In addition, IPW improves covariate balance and allows estimation of a marginal (population-average) effect that aligns with our study aims. Propensity scores were estimated using an unweighted logistic regression model [55], that included the covariates from the minimally sufficient adjustment set (age, race/ethnicity, BMI, smoking status, education, insurance status, age at menarche, and physical activity). This approach of variable selection into the propensity score model was chosen because prior studies suggest that including confounders and outcome-related covariates reduces bias more effectively than restricting the model to biological risk factors alone or variables that affect an exposure but not the outcome [56, 58, 59].

Stabilized IPWs were constructed using the marginal probability of endometriosis and applied to create a weighted pseudo-population in which measured covariates were balanced across exposure groups. Covariate balance before and after weighting was assessed using standardized mean differences (SMD), with values ≤0.10 indicating adequate balance [54]. In cases of residual imbalance, the propensity score model was modified by incorporating nonlinear terms in the relevant covariates (e.g., cubic terms) [54].

To obtain nationally representative marginal estimates, stabilized IPW were multiplied by NHANES survey weights to generate a combined weight that accounted for both confounding adjustments using IPW and the complex survey design (e.g., stratification and clustering) [55, 60]. The estimand of interest was the stratum-specific marginal association between endometriosis on early-onset CVD across levels of systemic inflammation on the odds-ratio scale. We fit weighted outcome models using survey-weighted logistic regression with the combined weight to estimate prevalence odds ratios and corresponding 95% confidence intervals. Since the CVD prevalence was low (3.1%) (Table 1), these odds ratios closely approximate prevalence ratios [61, 62]. Effect modification by systemic inflammation was assessed using stratified estimates and interaction terms in weighted models. Statistical significance was defined as a two-sided p < 0.05. All analyses were performed in R (version 4.3.2).

**Table 1.** Weighted Characteristics of U.S. Women Aged 20–54 Years by Endometriosis Status, NHANES 1999–2006.

| Characteristics | Total<br>(N =5,345) | Endometriosis |  | *p* |
| --- | --- | --- | --- | --- |
|  |  | Yes<br>(n = 370) | No<br>(n= 4,975) |  |
| **Age (years, Mean (S.E))** | 37.2 (0.2) | 40.7 (0.4) | 37.2 (0.2) | <0.001 |
| **Age at Menarche (years, Mean (S.E))<sup>1</sup>** | 12.6 (0.0) | 12.4 (0.1) | 12.6 (0.0) | 0.061 |
| **BMI, kg/m<sup>2</sup> (median, IQR)** | 26.5<br>(22.7, 32.3) | 26.9<br>(23.0, 32.1) | 26.5<br>(22.7, 32.3) | 0.646 |
| **Race, n (%)<sup>1</sup>** |  |  |  |  |
| Non-Hispanic White | 2,467 (46.2) | 254 (84.0) | 2,213 (66.9) | <0.001 |
| Non-Hispanic Black | 1,111 (20.8) | 65 (7.8) | 1,046 (12.8) |  |
| Hispanic | 1,527 (28.6) | 36 (4.2) | 1,491 (14.8) |  |
| Other | 240 (4.5) | 15 (4.0) | 225 (5.5) |  |
| **Education level, n (%)** |  |  |  |  |
| Less than High school | 1,291 (24.2) | 45 (9.9) | 1,246 (16.1) | <0.001 |
| High school grade/GED or equivalent | 1,193 (22.3) | 100 (31.6) | 1,093 (22.6) |  |
| More than High school | 2,861 (53.5) | 225 (58.5) | 2,636 (61.3) |  |
| **Health Insurance, n (%)** |  |  |  |  |
| Insured | 4,106 (76.8) | 315 (85.5) | 3,791 (80.7) | 0.058 |
| Uninsured | 1,239 (23.2) | 55 (14.5) | 1,184 (19.3) |  |
| **Cigarettes Smoking, n (%)** |  |  |  |  |
| Current | 1,165 (21.8) | 108 (32.9) | 1,057 (24.4) | 0.002 |
| Former | 870 (16.3) | 66 (18.3) | 804 (17.6) |  |
| Never | 3,310 (61.9) | 196 (48.8) | 3,114 (57.9) |  |
| **Physical Activity, n (%)** |  |  |  |  |
| Low | 1,320 (24.7) | 112 (32.7) | 1,208 (25.2) | 0.007 |
| Moderate | 2,977 (55.7) | 184 (50.7) | 2,793 (53.7) |  |
| High | 1,048 (19.6) | 74 (16.5) | 974 (21.1) |  |
| **Early-Onset CVD, n (%)** |  |  |  |  |
| Yes | 162 (3.0) | 30 (6.3) | 132 (2.8) | <0.001 |
| No | 5,183 (97.0) | 340 (93.7) | 4,843 (97.2) |  |
| **C-Reactive protein, mg/L (median, IQR)\*** | 2.3<br>(0.8, 5.7) | 3.1<br>(1.1, 6.4) | 2.3<br>(0.7, 5.6) | 0.002 |
| **C-Reactive protein, mg/L, n(%)** |  |  |  |  |
| Low/Moderate<br>(≤3 mg/L) | 2,574 (50.8) | 168 (49.5) | 2,406 (58.52) | 0.011 |
| High (>3mg/L) | 2,526 (49.3) | 188 (50.5) | 2,338 (49.5) |  |
| **Platelet ×10<sup>3</sup>/L, median (IQR)** | 283<br>(241, 332) | 286<br>(243, 330) | 282<br>(241, 332) | 0.786 |
| **Neutrophil ×10<sup>3</sup>/L, median (IQR)** | 4.3<br>(3.3, 5.5) | 4.3<br>(3.3, 5.5) | 4.3<br>(3.3, 5.3) | 0.782 |
| **Lymphocyte ×10<sup>3</sup>/L, median (IQR)** | 2.1<br>(1.7, 2.6) | 2.1<br>(1.7, 2.7) | 2.1<br>(1.7, 2.6) | 0.717 |
| **SII (median, IQR)\*** | 572<br>(418.5, 790.6) | 584.0<br>(418.2, 790.8) | 571.3<br>(418.6, 790.4) | 0.990 |
| **SII, n (%)** |  |  |  |  |
| Low (T1) | 1,587 (31.2) | 117 (33.3) | 1,470 (33.4) | 0.752 |
| Medium (T2) | 1,614 (31.8) | 108 (31.5) | 1,506 (33.5) |  |
| High (T3) | 1,880 (37.0) | 133 (35.2) | 1,747 (33.1) |  |

<sup>1</sup>n=unweighted frequencies; %= survey-weighted percentages
CRP (mg/L) was available for n = 5,100 participants and SII for n = 5,081; summaries for these biomarkers were calculated among participants with non-missing values.

## Assessment of propensity score overlap and covariate balance

We examined the distribution of estimated propensity scores to verify the overlap assumption. Figure 3 shows the kernel density plots of the propensity score among women with and without endometriosis. The two distributions demonstrated clear common support, with substantial overlap across most of the propensity score range. A small region of non-overlap was observed in the lower tail of the control distribution; however, overall overlap was sufficient to support IPW estimation. Stabilized inverse probability weights showed good overlap, with a mean near one and no evidence of extreme values (99th percentile = 1.97). Therefore, truncation or trimming of weights was not applied. Before weighting, several covariates showed substantial imbalance (unweighted SMDs up to 0.49). After application of IPW, balance improved markedly across all variables. Age showed some residual imbalance (SMD = 0.19) (Figure 4), likely reflecting a nonlinear association with endometriosis (Figure 5). To address this, age was modeled using restricted cubic splines in the propensity score model, which resulted in acceptable balance across all covariates (Supplementary Table 1).

**Figure 3:**
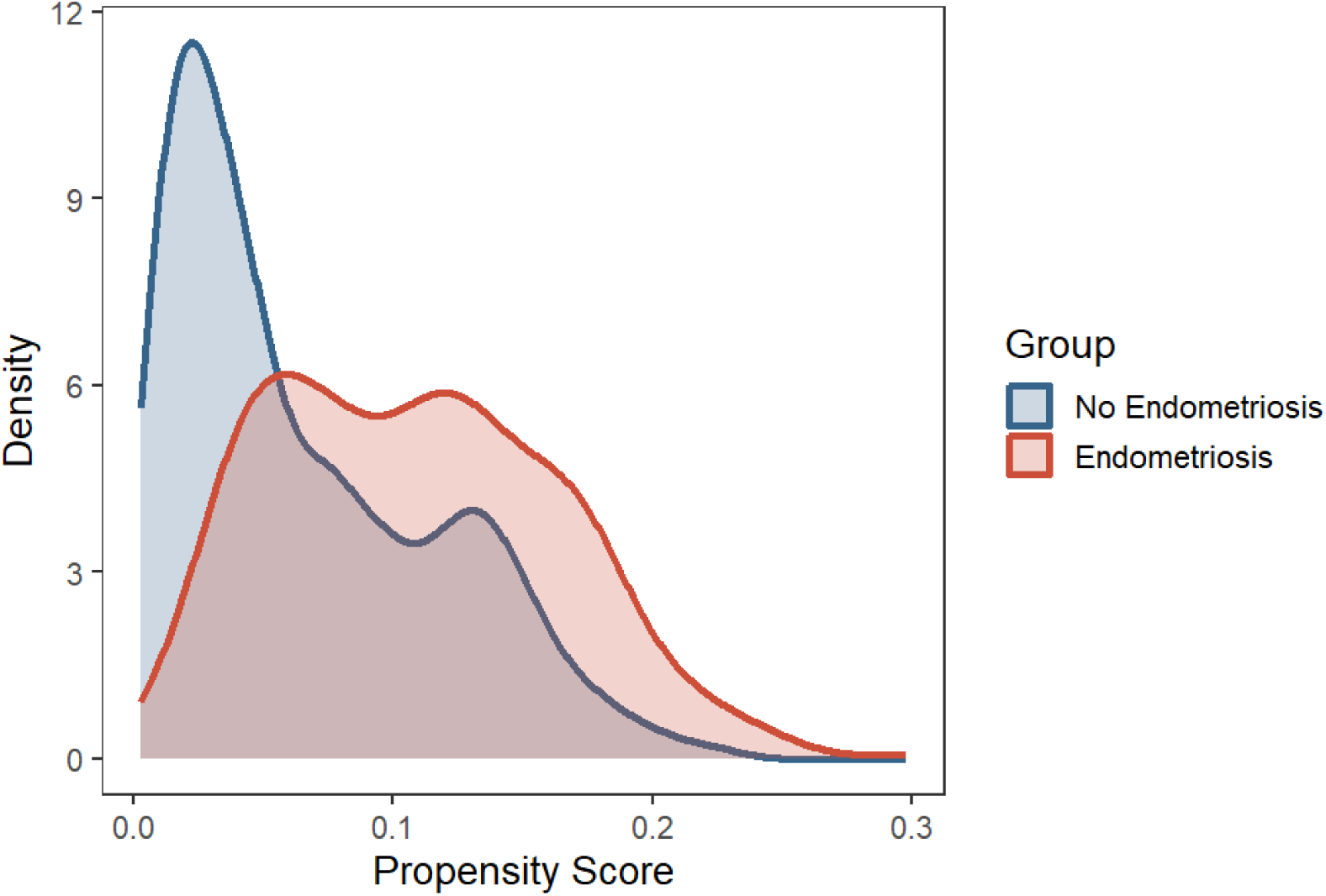
Propensity Score Overlap Between Women with and without Endometriosis prior to IPTW.

**Figure 4:**
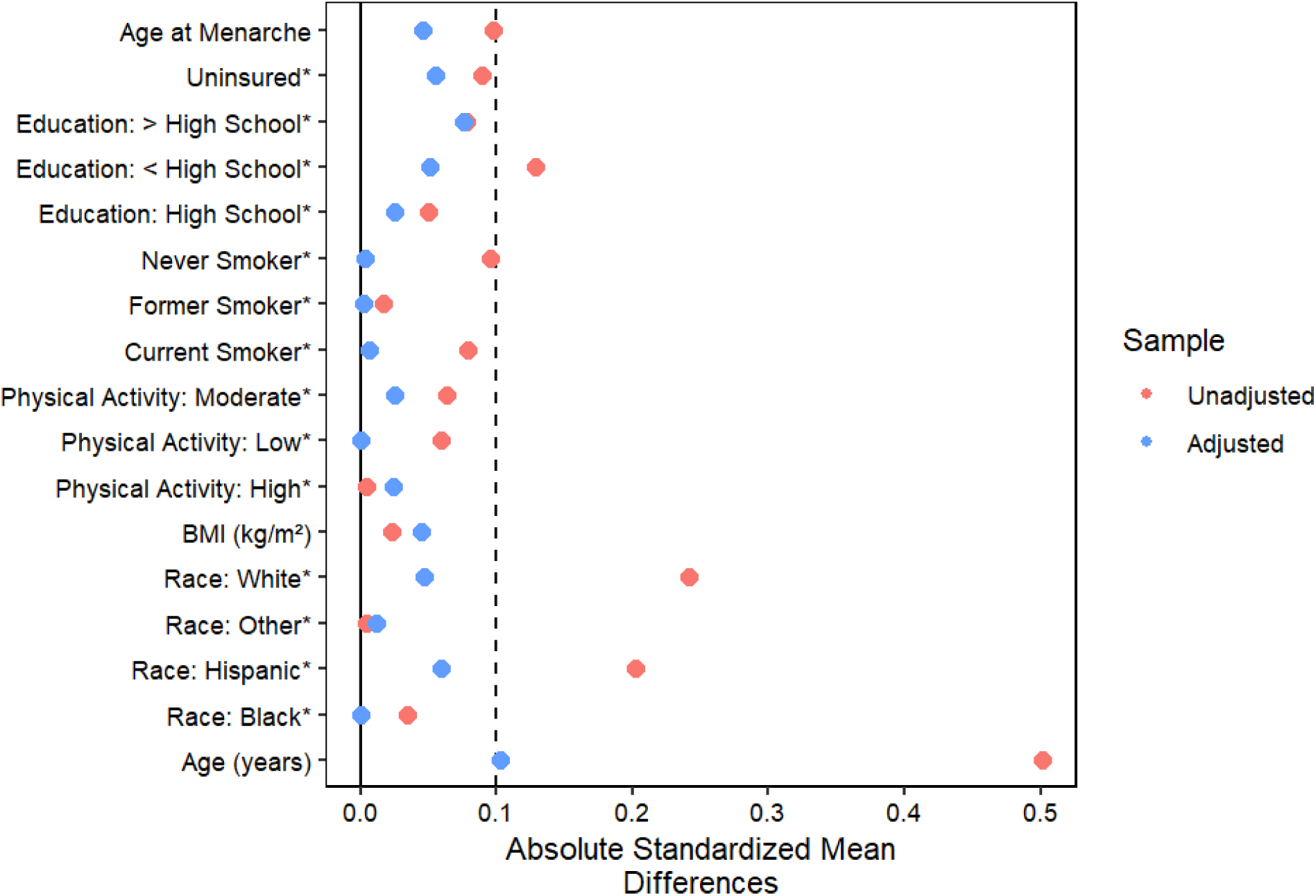
Covariate Balance Before and After IPTW Weighting.

**Figure 5:**
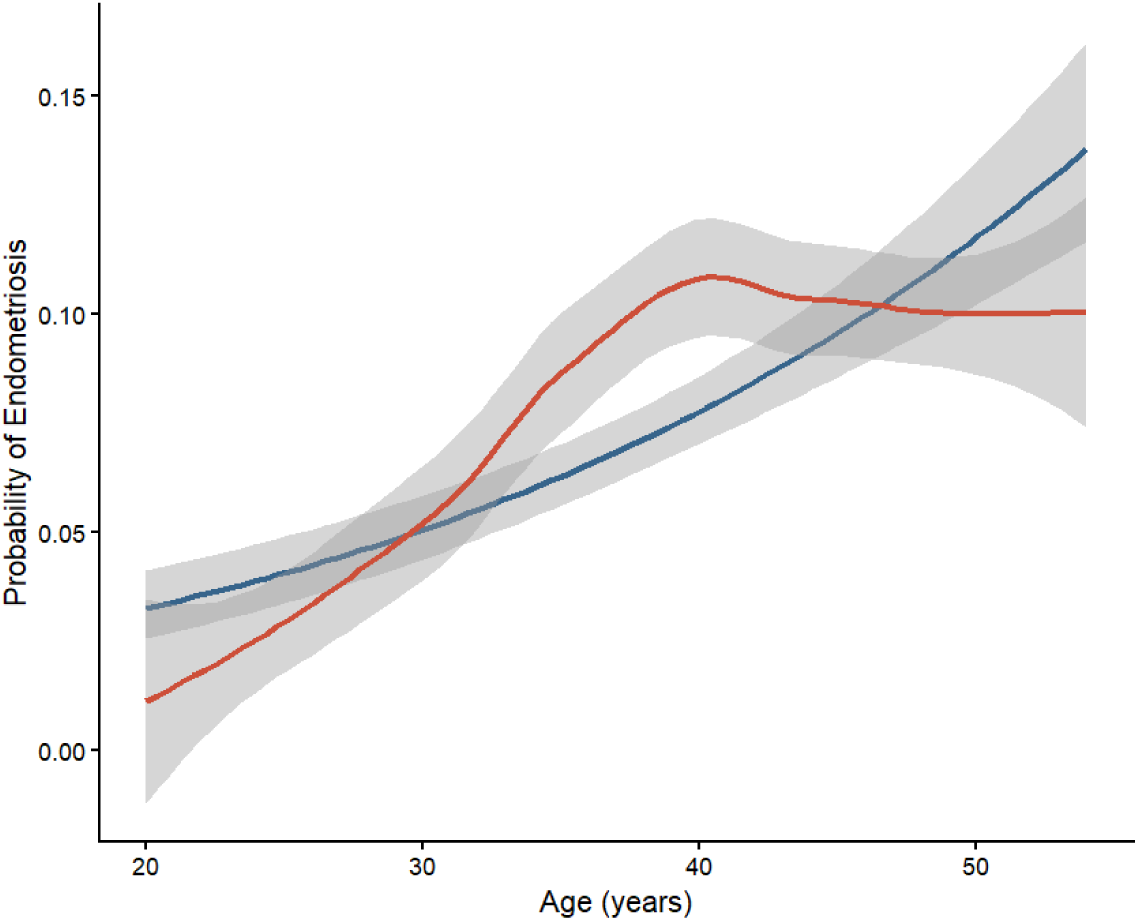
Non-linear Associations between Age and Endometriosis in the Propensity Score Model.

## Handling Missing Data

Overall missingness across study variables was low (≤5%), with the highest proportion observed for inflammatory markers (CRP and SII) (Supplementary Table 2). Missingness in CRP was not associated with endometriosis (p = 0.298) or CVD status (p = 0.302). Similarly, SII missingness was not associated with endometriosis (p= 0.079) or CVD (p= 0.347). Given the low level of missingness and absence of associations with key study variables, we conducted a complete-case analysis, restricting models to participants with non-missing data on the exposure, outcome, and all covariates. [63].

## Sensitivity Analysis

To evaluate the robustness of the IPW estimates, we conducted a survey-weighted logistic regression for the overall association between endometriosis and early-onset CVD. As a sensitivity analysis, participants with CRP values greater than 10 mg/L were excluded to reduce the influence of severe acute infection, ensuring findings represent chronic or low-grade inflammation rather than transient acute illness [64, 65]. Finally, in addition to SII tertiles in the primary analysis, we used log-transformed SII and survey-weighted median split to test for interaction on the multiplicative scale.

## Results

### 3.1. Baseline characteristics of the cohort

As shown in Table 1, our study included a total of 5,345 women aged 20–54 years, with a mean age of 37.2 ± 0.2 years. The survey-weighted prevalence of self-report endometriosis diagnosis was 6.9% and that for self-report early-onset CVD was 3.0%. The median (IQR) of CRP was 2.3 mg/L (0.8-5.7 mg/L) and SII was 572 (418.5, 790.6). The majority of the participants were non-Hispanic White women (46.2%) and had health insurance (76.8%). Approximately 53.5% had more than high school education, roughly one-quarter (21.8%) were current smokers and 19.6% reported high physical activity. The mean age at menarche was 12.6 ±0.0 years, while median (IQR) BMI 26.48 kg/m^2^ (22.71, 32.21 kg/m^2^).

Women with endometriosis were slightly older and more likely to be Non-Hispanic White and insured. They also had higher levels of CRP and SII and were more likely to report low physical activity, an earlier menarche, current smoking, and early-onset CVD compared with women without endometriosis. BMI was similar across groups.

## Association between Endometriosis and Early Onset CVD

In the unadjusted analysis, women with endometriosis had higher odds of early-onset CVD (POR = 2.35, 95% CI: 1.47-3.76). After IPW adjustment, the association remained significant, but was attenuated (aPOR = 1.91, 95% CI: 1.23-2.97) (Table 2).

**Table 2.** Associations Between Endometriosis and Early-Onset CVD.

| **Endometriosis** | **Crude**<br>**POR** | ***p*** | **Adjusted**<br>**POR\*** | ***p*** |
| --- | --- | --- | --- | --- |
| **Primary Analysis** | (95% CI) |  | (95% CI) |  |
| IPW | 2.35<br>(1.47-3.76) | <0.001 | 1.91<br>(1.23-2.97) | 0.005 |
| **Sensitivity Analysis** |  |  |  |  |
| Survey-weighted<br>logistic regression | 2.35<br>(1.47-3.76) | <0.001 | 1.96<br>(1.22-3.15) | 0.006 |

\* IPW-weighted Prevalence Odds Ratio
\*Adjusted for age, race/ethnicity, education level, physical activity, BMI, smoking status, age at menarche and insurance status

## Effect Modification by CRP

When stratified by CRP categories, endometriosis was associated with higher odds of early-onset CVD among women with CRP >3 mg/L (aPOR 2.33, 95% CI: 1.49–3.66), whereas no association was observed among those with CRP ≤3 mg/L (aPOR = 0.91, 95% CI: 0.35–2.31). However, the overall interaction test was not statistically significant (p for interaction = 0.086), suggesting that there was no strong statistical evidence of effect modification (Table 3).

**Table 3.** Association Between Endometriosis and Early-Onset CVD Stratified by CRP Category.

| CRP Category | POR<br>(95% CI) | p-value | p-interaction |
| --- | --- | --- | --- |
| Low/Moderate<br>($\leq 3$ mg/L) | 0.91<br>(0.35–2.31) | 0.836 | 0.086 |
| High (>3mg/L) | 2.33<br>(1.49-3.66) | <0.001 |  |

CVD events within CRP categories were: $\leq 3$ mg/L (7/168 Endo; 50/2,406 non-Endo) and $>3$ mg/L (22/188 Endo; 73/2,338 non-Endo).

## Effect Modification by SII

Table 4 shows the association between endometriosis and early-onset CVD across tertiles of SII. Among women in the lowest SII tertile, (aPOR = 1.25; 95% CI: 0.56–2.80) and highest SII tertile (aPOR = 1.72; 95% CI: 0.63–4.72), there was no evidence of an association between endometriosis and early-onset CVD. In contrast, women in the middle SII tertile had higher odds of early-onset CVD (aPOR = 2.92; 95% CI: 1.47–5.78). However, the global test for interaction was not statistically significant, also suggesting that there was no strong statistical evidence of effect modification (p for interaction = 0.288) in our study. (Table 4).

**Table 4.** Association Between Endometriosis and Early-Onset CVD Stratified by Tertiles of the Systemic Immune-Inflammation Index (SII)

| **SII Category** | **POR**<br>**(95% CI)** | **p-value** | **p-interaction** |
| --- | --- | --- | --- |
| **Primary Analysis** |  |  |  |
| Low (T1) | 1.25<br>(0.56–2.80) | 0.581 |  |
| Medium (T2) | 2.92<br>(1.47–5.78) | 0.002 | 0.288 |
| High (T3) | 1.72<br>(0.63–4.72) | 0.293 |  |
| **Sensitivity Analysis** |  |  |  |
| Continuous (log-SII)<sup>†</sup> | 1.97<br>(1.28-3.03) | 0.002 | 0.618 |
| **Median Split SII** |  |  |  |
| Low (<572) | 1.74<br>(0.98–3.10) | 0.059 | 0.566 |
| High (≥572) | 2.28<br>(1.18–4.42) | 0.015 |  |

CVD events within SII tertiles: T1 (9/117 Endo; 46/1470 non-Endo), T2 (14/108 Endo; 42/1506 non-Endo), T3 (7/133 Endo; 38/1747 non-Endo))
<sup>†</sup>POR evaluated at mean log-SII (6.39, SII≈597)

## Sensitivity Analyses

Using the survey-weighted logistic regression, women with endometriosis still had higher odds of early-onset CVD compared to women before (aPOR =2.35, 95% CI: 1.47–3.76) and after adjustment (aPOR =1.96, 95% CI: 1.22–3.15) (Table 2). Overall, the magnitude and direction of associations were consistent with those in the primary analysis. To assess the robustness of our findings to the influence of acute inflammation, we conducted a sensitivity analysis excluding participants with CRP values >10 mg/L as shown in Table 5. In this restricted sample (N = 4,286), endometriosis was not associated with early-onset CVD among women with CRP ≤3 mg/L (aPOR = 0.91, 95% CI: 0.35–2.31). Among women with CRP >3-10 mg/L, endometriosis remained associated with higher odds of early-onset CVD (aPOR = 2.25, 95% CI: 1.18–4.29) and the global interaction remained non-significant (p = 0.136). In sensitivity analyses, there was no significant interaction using log-transformed SII (p=0.618) and median split SII (p=0.566).

**Table 5.** Sensitivity Analysis: Association Between Endometriosis and Early-Onset CVD Stratified by CRP Category (CRP ≤ 10 mg/L)

| CRP Category | POR | p-value | p-interaction |
| --- | --- | --- | --- |
| Low/Moderate (≤3 mg/L) | 0.91<br>(0.35–2.31) | 0.836 | |
| High (>3–10 mg/L) | 2.25<br>(1.18–4.29) | 0.014 | 0.136 |

CVD events within CRP categories (excluding >10 mg/L) were: ≤ 3 mg/L (7/168 Endo; 50/2406 non-Endo) and >3–10 mg/L (13/124; 40/1588).

## Discussion

In this nationally representative study of U.S. women aged 20–54 years, we examined the association between endometriosis and early-onset CVD and examined heterogeneity by systemic inflammation measured by CRP and SII. First, we found that endometriosis was associated with higher odds of early-onset CVD. Second, the formal tests for interaction were not statistically significant, but the magnitude of the association between endometriosis and early-onset CVD differed across the strata of the inflammatory markers. Specifically, we found that the association was strongest among women with high CRP levels and those in the medium SII tertiles, whereas no association was observed among women with low/moderate CRP levels or in the low and high SII tertiles.

Our primary finding was that endometriosis was associated with higher odds of early-onset CVD. This observation is consistent with a growing body of evidence linking endometriosis to increased risk of overall CVD risk. For instance, prospective cohort studies and meta-analyses have reported modestly elevated risks of CVD events among women with endometriosis, including ischemic heart disease, stroke and composite CVD outcomes [19, 32, 33]. The biological plausibility linking endometriosis to CVD is well supported across multiple mechanisms. Genome-wide association studies have identified shared genetic loci between endometriosis and cardiovascular disease, suggesting common genetic susceptibility [66–68]. Beyond genetic overlap, endometriosis is associated with heightened oxidative stress and an atherogenic lipid profile, both of which are established contributors to atherosclerosis [69]. Additionally, endometriosis have been associated with elevated levels of high-density lipoprotein cholesterol (HDL-C) triglycerides, and pro-inflammatory cytokines including interleukin-6 (IL-6) and tumor necrosis factor-alpha (TNF-α) [70, 71]. All of which promote endothelial dysfunction, a key early step in atherosclerotic plaque formation [69]. Importantly, because endometriosis is typically diagnosed during the reproductive years, prolonged subclinical exposure to these factors may contribute to the development of early-onset CVD at younger ages than would be otherwise expected as noted in our study.

Our finding that the observed association between endometriosis and early-onset CVD was strongest among women with elevated CRP levels even after sensitivity analysis, may provide evidence to support the current hypothesis that the association between endometriosis and early-onset CVD may in part be driven by systemic inflammation [72]. Systemic inflammation creates an environment conducive to the growth and persistence of endometriosis lesions. In turn the ectopic endometriotic lesions become biologically active and capable of producing their own estrogen locally, which drives the growth of more lesions and fuels disease progression [73, 74]. This repetitive cycle leads to more inflammation which is an established risk factor for CVD. It is important to note that the formal test for interaction was not statistically significant. This may be due to lack of statistical power or true absence of statistical interaction. Therefore, our findings may only point to CRP as a potential marker associated with a higher prevalence of early-onset CVD in women with endometriosis but requires more studies to confirm this.

Finally, we found that the association between endometriosis and early-onset CVD was strongest among women in the medium SII tertile. SII been recognized as an important biomarker for CVD [75], and offers a more comprehensive assessment of systemic immune and inflammatory processes associated with endometriosis than CRP alone [76]. The strongest association between endometriosis and early-onset CVD was among women in moderate SII tertiles, but the formal interaction was not statistically significant. We hypothesize that this could be due to the following, first, the moderate SII tertiles may reflect chronic low-grade inflammation due to immune activation and inflammation, whereas the high SII tertiles may be as a result of acute infections and reflect a more transient state. Second, we may not have had sufficient power to detect meaningful effect modification. A prior study by Peng et al., [21] using NHANES data reported a nonlinear, S-shaped association between SII and endometriosis, with little association at lower SII levels and a stronger association emerging only after SII exceeded approximately 1100. In the present study, only 590 (11.0%) of our study population had SII values above 1100.

Taken together, our findings show an association between endometriosis and early-onset CVD and suggest that this association may vary by systemic inflammation. However, the non-significant interaction tests limit inferences about statistical interaction and as such our results should be interpreted cautiously and explored in future studies with sufficient power.

## Strengths and Limitations

This study has several notable strengths. First, we used data from NHANES, a nationally representative survey allowing our findings to generalize to U.S. women aged 20–54 years. We also applied inverse probability of weighting to address confounding which provides more reliable effect estimates. By evaluating two complementary markers of systemic inflammation CRP and SII, we were able to extend current literature on the association of systemic inflammation in the endometriosis and prevalent early-onset CVD. To our knowledge, this is the first study to formally examine the potential effect modification by systemic inflammation in the association between endometriosis and early-onset CVD.

However, several limitations should be considered. First, endometriosis and CVD were self-reported, which may introduce misclassification. Self-reported endometriosis is likely underdiagnosed, particularly among women with limited healthcare access. Because diagnosis depends on access to gynecologic care, misclassification may be differential with respect to socioeconomic status and healthcare utilization. If women with better access to care are more likely to receive both an endometriosis diagnosis and CVD diagnosis, this could bias associations away from the null. Second, the cross-sectional design eliminates the ability to establish temporality. Although we focused on early-onset CVD, the outcome reflects prevalent disease rather than incident events. As such, we cannot determine whether endometriosis or systemic inflammation preceded by early-onset CVD, resulted from it, or reflects shared underlying risk factors. Reverse causation remains possible. A prospective longitudinal design would be necessary to clarify the temporal sequence, which will be helpful in establishing causal inference. Third, systemic inflammatory markers were measured at a single time point. CRP and SII can fluctuate due to acute illness, circadian variation, medication use, or subclinical conditions. A single measurement may therefore misclassify inflammation levels. This misclassification is likely to be non-differential with respect to early-onset CVD and would generally bias effect estimate toward the null. Importantly, a single CRP value does not capture systemic inflammation. Fourth, although we used inverse probability weighting to address measured confounders, residual confounding due to reproductive history (e.g., parity, hysterectomy, oophorectomy), hormonal therapy use, disease severity, diet, medication use for endometriosis or CVD may influence both systemic inflammation and early-onset CVD which could obscure these associations. Additionally, smoking which is a major risk factor for CVD was categorized based on smoking status which is less reliable as it does not measure cumulative exposure to tobacco as does pack-years. While we used stabilized IPW combined with NHANES survey weights to reduce confounding by measured covariates which strengthens bias and confounding control relative to standard survey-weighted regression. However, combining multiple weighting can increase variance and reduce statistical precision, particularly for rare outcomes such as early-onset CVD and in stratified or interaction analyses. This may contribute to wider confidence intervals and non-significant interaction tests. Finally, the relatively small number of women with early-onset CVD limited statistical power to test for interaction, which may have reduced our ability to detect true effect modification. Although we observed patterns suggestive of heterogeneity by CRP and SII levels, our stratified analyses should be viewed as descriptive rather than causal and should be interpreted cautiously. Larger prospective studies will be needed to clarify these findings.

## Conclusion

In this nationally representative sample of U.S. women aged 20–54 years, we observed that the cross-sectional association between endometriosis and early-onset CVD was stronger at moderate to high levels of systemic inflammation, as reflected by SII and CRP respectively. Longitudinal studies with detailed clinical data are needed to clarify timing, mechanisms, and the role of inflammation in the progression from endometriosis to early cardiovascular disease.

## Data Availability

The data used in this study are publicly available from the National Health and Nutrition Examination Survey (NHANES), administered by the National Center for Health Statistics, Centers for Disease Control and Prevention. Data from the 1999-2006 survey cycles, including questionnaires, examination data, laboratory data, and documentation, are publicly available through the NHANES website.

https://wwwn.cdc.gov/nchs/nhanes/.

## Ethics approval and consent to participate

The NCHS Research Ethics Review Board approved the study protocol for the NHANES survey and all participants provided written informed consent.

## Consent for publication

Informed consent was obtained from all participants prior to their inclusion in the National Health and Nutrition Examination Survey (NHANES). All data were anonymized prior to public release and no individual patient consent for publication was required.

## Availability of data and materials

Publicly available Data from NHANES were used for this analysis and are available online at https://wwwn.cdc.gov/nchs/nhanes/Default.aspx.

## Competing interests

The authors declare no conflicts of interest related to this study.

## Funding

NC was supported National Institute on Alcohol Abuse and Alcoholism (NIAAA) grant K01AA029042. The content is solely the responsibility of the authors and does not represent the official views of the National Institutes of Health.

## CRediT authorship contribution statement

**Chidinma Oli (CO):** Conceptualization, Methodology, Formal analysis, Writing - Original Draft, Writing - Review & Editing, **Natalie Chichetto (NC):** Methodology, Writing - Review & Editing.

## Acknowledgements

We thank the NCHS Research Ethics Review Board and the participants who provided the data we used in this study.

## Notes

### Competing Interest Statement

The authors have declared no competing interest.

### Author Declarations

The study used only publicly available data from the National Health and Nutrition Examination Survey (NHANES), 1999-2006, conducted by the National Center for Health Statistics, Centers for Disease Control and Prevention. Public-use NHANES data and documentation are available from the NCHS NHANES website.

## References

1. Tsao CW, Aday AW, Almarzooq ZI, Anderson CA, Arora P, Avery CL, et al. Heart disease and stroke statistics—2023 update: a report from the American Heart Association. Circulation. 2023;147:e93.

2. Timmis A, Vardas P, Townsend N, Torbica A, Katus H, De Smedt D, et al. European Society of Cardiology: cardiovascular disease statistics 2021. European heart journal. 2022;43:716–99.

3. Mousavi I, Suffredini J, Virani SS, Ballantyne CM, Michos ED, Misra A, et al. Early-onset atherosclerotic cardiovascular disease. European Journal of Preventive Cardiology. 2025;32:100–12.

4. Bucholz EM, Strait KM, Dreyer RP, Lindau ST, D’Onofrio G, Geda M, et al. Editor’s choice-sex differences in young patients with acute myocardial infarction: a VIRGO study analysis. European Heart Journal: Acute Cardiovascular Care. 2017;6:610–22.

5. Collet J-P, Zeitouni M, Procopi N, Hulot J-S, Silvain J, Kerneis M, et al. Long-term evolution of premature coronary artery disease. Journal of the American college of cardiology. 2019;74:1868–78.

6. Steen Carlsson K, Nilsson K, Wolden ML, Faurby M. Economic burden of atherosclerotic cardiovascular disease: a matched case–control study in more than 450,000 Swedish individuals. BMC Cardiovascular Disorders. 2023;23:483.

7. Williams KJ. Eradicating atherosclerotic events by targeting early subclinical disease: it is time to retire the therapeutic paradigm of too much, too late. Arteriosclerosis, Thrombosis, and Vascular Biology. 2024;44:48–64.

8. Liu X, Wu Y, Li F, Qi X, Niu L, Wu Y, et al. Global Burden of Early-Onset Ischemic Heart Disease, 1990 to 2019. JACC: Advances. 2025;4:101466.

9. Rajendran A, Minhas AS, Kazzi B, Varma B, Choi E, Thakkar A, et al. Sex-specific differences in cardiovascular risk factors and implications for cardiovascular disease prevention in women. Atherosclerosis. 2023;384:117269.

10. Bancks MP, Akhabue E, Rana JS, Reis JP, Schreiner PJ, Yano Y, et al. Sex differences in cardiovascular risk factors before and after the development of type 2 diabetes and risk for incident cardiovascular disease. Diabetes Research and Clinical Practice. 2020;166:108334.

11. Nguyen AH, Hurwitz M, Sullivan SA, Saad A, Kennedy JL, Sharma G. Update on sex specific risk factors in cardiovascular disease. Frontiers in cardiovascular medicine. 2024;11:1352675.

12. Fan J, Chen J, Wang L, Yuan C, Liu X, Wang J. The global burden and attributable risk factor analysis of cardiovascular disease between females and males, 1990–2021: findings from the 2021 global burden of disease study. International Journal of Surgery. 2026;112:239–49.

13. Mu F, Rich-Edwards J, Rimm EB, Spiegelman D, Missmer SA. Endometriosis and risk of coronary heart disease. Circulation: Cardiovascular Quality and Outcomes. 2016;9:257–64.

14. Okoth K, Wang J, Zemedikun D, Thomas GN, Nirantharakumar K, Adderley N. Risk of cardiovascular outcomes among women with endometriosis in the United Kingdom: a retrospective matched cohort study. BJOG: An International Journal of Obstetrics & Gynaecology. 2021;128:1598–609.

15. Taylor HS, Kotlyar AM, Flores VA. Endometriosis is a chronic systemic disease: clinical challenges and novel innovations. Lancet. 2021;397:839–52.

16. Chiang H-J, Lan K-C, Yang Y-H, Chiang JY, Kung F-T, Huang F-J, et al. Risk of major adverse cardiovascular and cerebrovascular events in Taiwanese women with endometriosis. Journal of the Formosan Medical Association. 2021;120:327–36.

17. Wei C-H, Chang R, Wan YH, Hung Y-M, Wei JC-C. Endometriosis and new-onset coronary artery disease in Taiwan: a nationwide population-based study. Frontiers in medicine. 2021;8:619664.

18. Simancas-Racines D, Jiménez-Flores E, Montalvan M, Horowitz R, Araujo V, Reytor-González C. Endometriosis as a Systemic and Complex Disease: Toward Phenotype-Based Classification and Personalized Therapy. International Journal of Molecular Sciences. 2026;27:908.

19. Mu F, Rich-Edwards J, Rimm EB, Spiegelman D, Missmer SA. Endometriosis and risk of coronary heart disease. Circulation: Cardiovascular Quality and Outcomes. 2016;9:257–64.

20. Rafique S, Decherney A. Medical Management of Endometriosis. Clinical Obstetrics and Gynecology. 2017;60:485–96.

21. Peng L, Luo X, Cao B, Wang X. Exploring the link: systemic immune-inflammation index as a marker in endometriosis—insights from the NHANES 2001–2006 cross-sectional study. PloS one. 2024;19:e0304591.

22. Rafi U, Ahmad S, Bokhari SS, Iqbal MA, Zia A, Khan MA, et al. Association of inflammatory markers/cytokines with cardiovascular risk manifestation in patients with endometriosis. Mediators of inflammation. 2021;2021:3425560.

23. Vallvé-Juanico J, Houshdaran S, Giudice LC. The endometrial immune environment of women with endometriosis. Human reproduction update. 2019;25:565–92.

24. Xu X, Cheng H, Ye X, Chang X. Association Between the C-Reactive Protein–Albumin–Lymphocyte Index and Endometriosis: Insights from the 1999–2006 National Health and Nutrition Examination Survey. International Journal of Women’s Health. 2025;:4267–80.

25. Rathod S, Shanoo A, Acharya N. Endometriosis: a comprehensive exploration of inflammatory mechanisms and fertility implications. Cureus. 2024;16.

26. Smyk JM, Danielecka Z, Kotowska M, Zawadka M, Andruszkiewicz P, Grąt M, et al. Cardiovascular risks and endothelial dysfunction in reproductive-age women with endometriosis. Scientific Reports. 2024;14:24127.

27. Tan J, Taskin O, Iews M, Lee AJ, Kan A, Rowe T, et al. Atherosclerotic cardiovascular disease in women with endometriosis: a systematic review of risk factors and prospects for early surveillance. Reproductive biomedicine online. 2019;39:1007–16.

28. Davignon J, Ganz P. Role of endothelial dysfunction in atherosclerosis. Circulation. 2004;109 23_suppl_1:III–27.

29. Devaraj S, Kumaresan PR, Jialal I. C-reactive protein induces release of both endothelial microparticles and circulating endothelial cells in vitro and in vivo: further evidence of endothelial dysfunction. Clinical chemistry. 2011;57:1757–61.

30. Xu S, Ilyas I, Little PJ, Li H, Kamato D, Zheng X, et al. Endothelial dysfunction in atherosclerotic cardiovascular diseases and beyond: from mechanism to pharmacotherapies. Pharmacological reviews. 2021;73:924–67.

31. As-Sanie S, Mackenzie SC, Morrison L, Schrepf A, Zondervan KT, Horne AW, et al. Endometriosis: a review. Jama. 2025.

32. Blom JN, Velez MP, McClintock C, Shellenberger J, Pudwell J, Brogly SB, et al. Endometriosis and cardiovascular disease: a population-based cohort study. Canadian Medical Association Open Access Journal. 2023;11:E227–36.

33. Parsa S, Noroozpoor R, Dehghanbanadaki H, Khateri S, Moradi Y. Endometriosis and risk of cardiovascular disease: a systematic review and meta-analysis. BMC Public Health. 2025;25:1–8.

34. Machairiotis N, Vasilakaki S, Thomakos N. Inflammatory mediators and pain in endometriosis: a systematic review. Biomedicines. 2021;9:54.

35. Ustundag Y, Huysal K, Gecgel SK, Unal D. Relationship between C-reactive protein, systemic immune-inflammation index, and routine hemogram-related inflammatory markers in low-grade inflammation. Int J Med Biochem. 2018;1:24–8.

36. Youkhana S, Odusanmi SS, Ihuchukwu BK, Ezekiel M, Enyeneokpon E, Cumaaran C, et al. Glucagon-Like Peptide-1 (GLP-1) Receptor Agonist Use and Inflammatory Markers Among US Adults: A National Health and Nutrition Examination Survey (NHANES)-Based Analysis. Cureus. 2025;17:e90964.

37. Herington JL, Bruner-Tran KL, Lucas JA, Osteen KG. Immune interactions in endometriosis. Expert review of clinical immunology. 2011;7:611–26.

38. Hu B, Yang X-R, Xu Y, Sun Y-F, Sun C, Guo W, et al. Systemic immune-inflammation index predicts prognosis of patients after curative resection for hepatocellular carcinoma. Clinical Cancer Research. 2014;20:6212–22.

39. Curtin LR, Mohadjer LK, Dohrmann SM, Montaquila JM, Kruszan-Moran D, Mirel LB, et al. The National Health and Nutrition Examination Survey: Sample Design, 1999-2006. Vital and health statistics Series 2, Data evaluation and methods research. 2012;:1–39.

40. Shafrir A, Wise L, Palmer J, Shuaib Z, Katuska L, Vinayak P, et al. Validity of self-reported endometriosis: a comparison across four cohorts. Human Reproduction. 2021;36:1268–78.

41. Lee AJ, Sanchez D, Reyes-Dumeyer D, Brickman AM, Lantigua RA, Vardarajan BN, et al. Reliability and validity of self-reported vascular risk factors: hypertension, diabetes, and heart disease, in a multi-ethnic community based study of aging and dementia. Journal of Alzheimer’s Disease. 2023;95:275–85.

42. Sabanayagam C, He F, Chee ML, Cheng C-Y. Validity and reliability of Self-Reported prevalent and incident cardiovascular disease among Asian adults. Journal of Cardiovascular Development and Disease. 2024;11:350.

43. McQuillan GM, McLean JE, Chiappa M, Lukacs SL. National Health and nutrition examination survey biospecimen program: NHANES III (1988–1994) and NHANES 1999–2014. 2015.

44. Pearson TA, Mensah GA, Alexander RW, Anderson JL, Cannon III RO, Criqui M, et al. Markers of inflammation and cardiovascular disease: application to clinical and public health practice: a statement for healthcare professionals from the Centers for Disease Control and Prevention and the American Heart Association. circulation. 2003;107:499–511.

45. Ridker PM. C-reactive protein: a simple test to help predict risk of heart attack and stroke. Circulation. 2003;108:e81–5.

46. Casagrande SS, Lawrence JM. Cardiovascular disease risk factors and their associations with inflammation among US adolescents: NHANES, 2015 to March 2020. BMJ Open Diabetes Research and Care. 2024;12:e004148.

47. Goldstein A, Cohen S. Self-report symptom-based endometriosis prediction using machine learning. Scientific Reports. 2023;13:5499.

48. Liu X, An R, Wang G. Integrating inflammatory biomarkers and demographic variables with machine learning to predict endometriosis risk. Scientific Reports. 2025;15:42415.

49. Peng M, Hou F, Cheng Z, Shen T, Liu K, Zhao C, et al. Prediction of cardiovascular disease risk based on major contributing features. Scientific Reports. 2023;13:4778.

50. Shafrir AL, Farland LV, Shah DK, Harris HR, Kvaskoff M, Zondervan K, et al. Risk for and consequences of endometriosis: A critical epidemiologic review. Best Practice & Research Clinical Obstetrics & Gynaecology. 2018;51:1–15. 10.1016/j.bpobgyn.2018.06.001.

51. Klasson CL, Sadhir S, Pontzer H. Daily physical activity is negatively associated with thyroid hormone levels, inflammation, and immune system markers among men and women in the NHANES dataset. PLoS One. 2022;17:e0270221.

52. Trexler ET, Eagle DE, Pontzer H. Physical activity and diet pattern do not mitigate C-reactive protein increases associated with oral contraceptive use. PLoS One. 2025;20:e0319928.

53. National Center for Health Statistics. Weighting: National Health and Nutrition Examination Survey. 2025. https://wwwn.cdc.gov/nchs/nhanes/tutorials/weighting.aspx#print. Accessed 8 May 2025.

54. Austin PC, Stuart EA. Moving towards best practice when using inverse probability of treatment weighting (IPTW) using the propensity score to estimate causal treatment effects in observational studies. Statistics in medicine. 2015;34:3661–79.

55. DuGoff EH, Schuler M, Stuart EA. Generalizing observational study results: applying propensity score methods to complex surveys. Health services research. 2014;49:284–303.

56. Austin PC. An introduction to propensity score methods for reducing the effects of confounding in observational studies. Multivariate behavioral research. 2011;46:399–424.

57. Valojerdi AE, Janani L. A brief guide to propensity score analysis. Medical journal of the Islamic Republic of Iran. 2018;32:122.

58. Austin PC. The performance of different propensity-score methods for estimating relative risks. Journal of Clinical Epidemiology. 2008;61:537–45. 10.1016/j.jclinepi.2007.07.011.

59. Brookhart MA, Schneeweiss S, Rothman KJ, Glynn RJ, Avorn J, Stürmer T. Variable selection for propensity score models. American journal of epidemiology. 2006;163:1149–56.

60. Cook BL, McGuire TG, Meara E, Zaslavsky AM. Adjusting for health status in non-linear models of health care disparities. Health Services and Outcomes Research Methodology. 2009;9:1–21.

61. Barros AJ, Hirakata VN. Alternatives for logistic regression in cross-sectional studies: an empirical comparison of models that directly estimate the prevalence ratio. BMC medical research methodology. 2003;3:21.

62. Martinez BAF, Leotti VB, Silva G de S e, Nunes LN, Machado G, Corbellini LG. Odds ratio or prevalence ratio? An overview of reported statistical methods and appropriateness of interpretations in cross-sectional studies with dichotomous outcomes in veterinary medicine. Frontiers in veterinary science. 2017;4:193.

63. Heymans MW, Twisk JWR. Handling missing data in clinical research. Journal of Clinical Epidemiology. 2022;151:185–8. 10.1016/j.jclinepi.2022.08.016.

64. Ridker PM. From C-reactive protein to interleukin-6 to interleukin-1: moving upstream to identify novel targets for atheroprotection. Circulation research. 2016;118:145–56.

65. Wu D, Chen G, Lan Y, Chen S, Ding X, Wei C, et al. Measurement of cumulative high-sensitivity C-reactive protein and monocyte to high-density lipoprotein ratio in the risk prediction of type 2 diabetes: a prospective cohort study. Journal of Translational Medicine. 2024;22:110.

66. Kalaitzopoulos DR, Samartzis N, Kolovos GN, Mareti E, Samartzis EP, Eberhard M, et al. Treatment of endometriosis: a review with comparison of 8 guidelines. BMC women’s health. 2021;21:397.

67. Nyholt DR, Low S-K, Anderson CA, Painter JN, Uno S, Morris AP, et al. Genome-wide association meta-analysis identifies new endometriosis risk loci. Nature genetics. 2012;44:1355–9.

68. Vazgiourakis VM, Zervou MI, Papageorgiou L, Chaniotis D, Spandidos DA, Vlachakis D, et al. Association of endometriosis with cardiovascular disease: Genetic aspects. International Journal of Molecular Medicine. 2023;51:29.

69. Marchandot B, Faller E, Akladios C, Matsushita K, Bäck M, Jesel L, et al. Fostering cardio-endometriosis: a call to action for a comprehensive understanding of cardiovascular disease in endometriosis. European Journal of Preventive Cardiology. 2024;31:1574–82.

70. Duan Y, Kong F, Ding Z, Peng Y, Chen A, Yao Y. Impact of the lipid–inflammation axis on endometriosis risk: a multicenter case–control study using mediation analysis. Frontiers in Endocrinology. 2025;16. 10.3389/fendo.2025.1661264.

71. Hameed H, Qadir A, Hussain T, Qurshi W, Inayat A, Baloch NS. Biochemical Profiling of Inflammatory and Metabolic Markers in Reproductive-Age Women with Endometriosis. Pakistan Journal of Medical and Health Sciences. 2023. 10.53350/pjmhs020231712356.

72. Poeta do Couto C, Policiano C, Pinto FJ, Brito D, Caldeira D. Endometriosis and cardiovascular disease: A systematic review and meta-analysis. Maturitas. 2023;171:45–52. 10.1016/j.maturitas.2023.04.001.

73. Bulun SE, Monsavais D, Pavone ME, Dyson M, Xue Q, Attar E, et al. Role of estrogen receptor-β in endometriosis. Thieme Medical Publishers; 2012. p. 39–45.

74. Mick I, Freger SM, van Keizerswaard J, Gholiof M, Leonardi M. Comprehensive endometriosis care: a modern multimodal approach for the treatment of pelvic pain and endometriosis. Therapeutic advances in reproductive health. 2024;18:26334941241277759.

75. Xu J-P, Zeng R-X, Zhang Y-Z, Lin S-S, Tan J-W, Zhu H-Y, et al. Systemic inflammation markers and the prevalence of hypertension: a NHANES cross-sectional study. Hypertension Research. 2023;46:1009–19.

76. Zhao J, Zeng L, Liang G, Dou Y, Zhou G, Pan J, et al. Higher systemic immune-inflammation index is associated with sarcopenia in individuals aged 18–59 years: a population-based study. Scientific Reports. 2023;13:22156.

