## Supplementary Tables for "Endometriosis and Early-Onset Cardiovascular Disease Across Levels of Systemic Inflammation: A Cross-Sectional Study of NHANES 1999–2006"

**Supplementary Table 1. Standardized mean differences (SMD) of baseline covariates comparing women with and without endometriosis before and after inverse probability weighting (N=5,345)**

| Covariate | SMD Before Weighting | SMD After Weighting |
| --- | --- | --- |
| Age (years) | 0.50 | 0.10 |
| Race/Ethnicity |  |  |
| Black | -0.03 | -0.00 |
| Hispanic | -0.20 | -0.06 |
| Other | -0.00 | 0.01 |
| White | 0.24 | 0.05 |
| BMI (kg/m²) | -0.02 | -0.04 |
| Physical Activity |  |  |
| Low | 0.06 | -0.00 |
| Moderate | -0.06 | -0.03 |
| High | 0.00 | 0.02 |
| Smoking Status |  |  |
| Current Smoker | 0.08 | -0.01 |
| Former Smoker | 0.02 | 0.00 |
| Never Smoker | -0.10 | -0.00 |
| Education Level |  |  |
| Less Than High School | -0.13 | -0.05 |
| High School Diploma | 0.05 | -0.03 |
| More Than High School | 0.08 | 0.08 |
| Insurance Status |  |  |
| Uninsured | -0.09 | -0.06 |
| Age at Menarche (Years) | -0.10 | -0.05 |

**Supplementary Table 2: Missing Variables in NHANES 1999–2006 for U.S women aged 1996-2006 across covariates in the eligible study population (n = 5,575).**

| Variable | Total  N | Missing (n) | Missing (%) |
| --- | --- | --- | --- |
| Age | 5,575 | 0 | 0 |
| Race/Ethnicity | 5,575 | 0 | 0 |
| BMI | 5,517 | 58 | 1.04 |
| Smoking Status | 5,571 | 4 | 0.07 |
| Education Level | 5,570 | 5 | 0.09 |
| Insurance Status | 5,528 | 47 | 0.85 |
| Physical Activity | 5,572 | 3 | 0.05 |
| Age at Menarche | 5,477 | 98 | 1.76 |
| Endometriosis | 5,557 | 18 | 0.32 |
| Early-onset CVD | 5,564 | 11 | 0.20 |
| CRP | 5,309 | 266 | 4.79 |
| SII | 5,292 | 283 | 5.09 |
